# Methadone overdoses increased 48% during the COVID-19 epidemic

**DOI:** 10.1101/2022.04.14.22273870

**Authors:** Daniel E. Kaufman, Amy L. Kennalley, Kenneth L McCall, Brian J. Piper

## Abstract

**Background:** The United States (US) is in the middle of an opioid overdose epidemic that has spanned over two decades and continues to escalate. Methadone is long-acting full opioid agonist which has been approved to treat opioid use disorder (OUD). Methadone can cause respiratory depression that may result in mortality. The restrictions on methadone availability including take-home dosing were loosened during the COVID-19 pandemic although there have been concerns about the high street value of diverted methadone. This report examined how fatal overdoses involving methadone have changed over the past two-decades including during the pandemic.

**Methods:** The Center for Disease Control and Prevention’s Wide-ranging Online Data for Epidemiologic Research (WONDER) database, which draws data from death certificates, was used to find the unintentional methadone related overdose death rate from 1999-2020. Unintentional methadone deaths were defined using the International Statistical Classification of Diseases (ICD), 10^th^ revision codes: X40-44 with only data which was coded for methadone (T40.3). Data from the Automation of Reports and Consolidated Orders System (ARCOS) on methadone overall use, narcotic treatment programs use, and pain management use was gathered for all states, including the District of Columbia, for 2020 and corrected for population.

**Results:** There have been dynamic changes over the past two-decades in overdoses involving methadone. Overdoses increased from 1999 (0.9/million) to 2007 (15.9) and declined until 2019 (6.5). Overdoses in 2020 (9.6) were 48.1% higher than in 2019 (*t*(50) = 3.05, *p* < .005).

The correlations between overall methadone use (*r*(49) = +0.75, *p* < 0.001), and narcotic treatment program use (*r*(49) = +0.77, *p* < 0.001) were positive, strong, and statistically significant. However, methadone use for pain treatment was not associated with overdoses (*r*(49) = -.08, *p* = .57).

**Conclusions:** Overdoses involving methadone significantly increased by 48.1% in 2020 relative to 2019. This mortality increase is much larger than the 5.3% elevation in calls involving methadone reported to poison control centers in the year following the March 16, 2020 loosening of methadone take-home regulations. Policy changes that were implemented following the COVID-19 pandemic involving methadone may warrant reconsideration.

---

The United States (US) is in the middle of an opioid overdose epidemic that has spanned over two decades and continues to escalate [1]. Methadone is long-acting mu receptor full opioid agonist approved to treat opioid use disorder (OUD) [2]. Methadone maintenance reduces the illicit use of heroin, death rates and criminality associated with heroin use, and allows patients to improve their health and social productivity [2]. Methadone induced respiratory depression may lead to mortality. The restrictions on methadone availability including take-home dosing were relaxed during the COVID-19 pandemic although there have been concerns about the high street value of diverted methadone [3]. Calls involving methadone to poison control centers increased 5.3% during the pandemic [4]. This report examined how fatal overdoses involving methadone have changed over the past two-decades including following the pandemic.

The Center for Disease Control and Prevention’s (CDC) Wide-ranging Online Data for Epidemiologic Research (WONDER) database [5], which draws data from death certificates, was used to find the unintentional methadone related overdose death rate from 1999-2020. Unintentional methadone deaths were defined using the International Statistical Classification of Diseases (ICD), 10^th^ revision codes: X40-44 with only data which was coded for methadone (T40.3). States with overdoses that were so low that results were suppressed (32.5% of values), were coded as 0. Data from the Automation of Reports and Consolidated Orders System (ARCOS) on methadone overall use, narcotic treatment programs use, and pain management (including pharmacies, hospitals, and mid-level practitioners) use was gathered for all states, including the District of Columbia, for 2020 and corrected for population. Procedures were approved as exempt by the Geisinger and University of New England IRBs. Data-analysis was completed with Systat and Figures were prepared with GraphPad Prism.

There have been dynamic changes over the past two-decades in overdoses involving methadone. Overdoses increased from 1999 to 2007 and declined until 2019. Overdoses in 2020 during the pandemic were 48.1% higher than in 2019 (*t*(50) = 3.05, *p* < .005, Figure 1A).Among the top ten states (Rhode Island = 4.64 / 100,000, Washington DC = 4.07, Delaware = 3.34, Connecticut = 3.12, New Mexico = 2.66, West Virginia = 2.07, New York = 2.06, Maine = 1.85, Illinois = 1.66, Massachusetts = 1.54), nine were located in the eastern US (Supplemental Table 1). Further, the correlations between overall methadone use (*r*(49) = +0.75, *p* < 0.001, not shown), and narcotic treatment program use (*r*(49) = +0.77, *p* < 0.001, Figure 1B) was positive, strong, and statistically significant. However, methadone for pain treatment was not associated with overdoses (Figure 1C).

**Figure 1.**
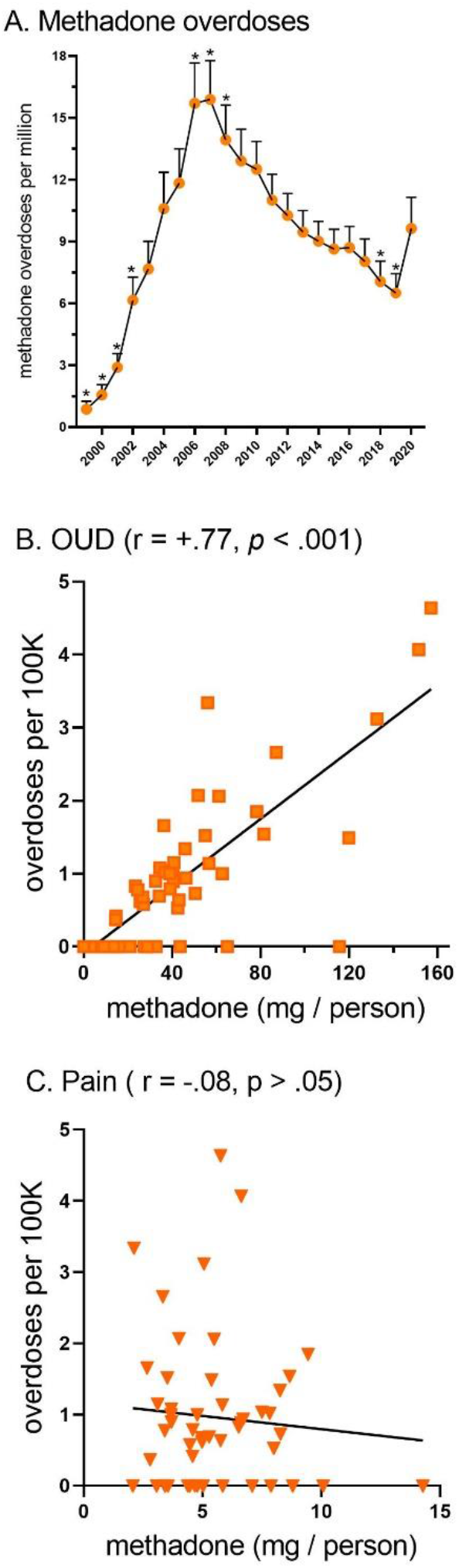
Dynamic changes in overdoses involving methadone as reported to the Center for Disease Control and Prevention’s Wide-ranging Online Data for Epidemiologic Research (WONDER) database (A, * *p* < .05 versus 2020). Correlations between per capita methadone distribution for Opioid Use Disorder (OUD, B) and pain (C) treatment in 2020 as reported to the Drug Enforcement Administration’s Automated Reports and Consolidated Orders System.

Overdoses involving methadone significantly increased by 48.1% in 2020 relative to 2019. This increase is consistent with but also much larger than the 5.3% elevation in calls involving methadone reported to poison control centers in the year following the March 16, 2020 loosening of methadone take-home regulations.

It is also important to recognize that death determination is not a simple process and there are substantial state-level differences in reporting quality [6] which could non-homogeneously impact the detection of a drug with a long half-life like methadone relative to other more obscure substances (fentanyl analogues or xylazine). Although methadone fatalities are much less common than those involving heroin or fentanyl, these findings of increased overdoses involving methadone may warrant reconsideration of the COVID-19 loosening of methadone policies.

## Supporting information

methadone_overdoses_WONDER

methadone_distribution_DEA_ARCOS

## Data Availability

Raw data for overdoses may be found at: https://wonder.cdc.gov/
Raw data for methadone distribution may be found at: https://www.deadiversion.usdoj.gov/arcos/retail_drug_summary/

https://www.deadiversion.usdoj.gov/arcos/retail_drug_summary/

## Acknowledgements

BJP is supported by the Health Resources Services Administration (D34HP31025). The National Institute of Environmental Health Sciences (T32 ES007060-31A1) provided software used for figure construction.

## Conflicts of Interest

BJP was part of an osteoarthritis research team supported by Pfizer and Eli Lilly from 2019-2021. The other authors have no relevant disclosures.

---

**Supplemental Table 1.** States ranked for methadone overdoses per 100,000 population in 2020 as reported to the Center for Disease Control and Prevention’s (CDC) Wide-ranging Online Data for Epidemiologic Research (WONDER). Sixteen states (Alaska, Arkansas, Hawaii, Iowa, Idaho, Kansas, Mississippi, Montana, Nebraska, New Hampshire, North Dakota, Oklahoma, South Dakota, Utah, Vermont, and Wyoming) did not report any methadone overdoses.

| <u>Rank</u> | <u>State</u> | <u>Overdoses</u> | <u>Rank</u> | <u>State</u> | <u>Overdoses</u> |
| --- | --- | --- | --- | --- | --- |
| 1 | Rhode Island | 4.64 | 19 | Arizona | 1.00 |
| 2 | Washington DC | 4.07 | 20 | Virginia | 1.00 |
| 3 | Delaware | 3.34 | 21 | Kentucky | 0.94 |
| 4 | Connecticut | 3.12 | 22 | Minnesota | 0.90 |
| 5 | New Mexico | 2.66 | 23 | North Carolina | 0.89 |
| 6 | West Virginia | 2.07 | 24 | Florida | 0.83 |
| 7 | New York | 2.06 | 25 | South Carolina | 0.79 |
| 8 | Maine | 1.85 | 26 | Tennessee | 0.78 |
| 9 | Illinois | 1.66 | 27 | Oregon | 0.73 |
| 10 | Massachusetts | 1.54 | 28 | Georgia | 0.69 |
| 11 | New Jersey | 1.52 | 29 | Ohio | 0.68 |
| 12 | Maryland | 1.49 | 30 | Indiana | 0.64 |
| 13 | Washington | 1.34 | 31 | California | 0.62 |
| 14 | Wisconsin | 1.15 | 32 | Louisiana | 0.58 |
| 15 | Pennsylvania | 1.14 | 33 | Alabama | 0.53 |
| 16 | Colorado | 1.08 | 34 | Missouri | 0.42 |
| 17 | Michigan | 1.04 | 35 | Texas | 0.37 |
| 18 | Nevada | 1.02 |  |  |  |

## Notes

### Funding Statement

This study did not receive any external funding. BJP is supported by the Health Resources Services Administration (D34HP31025). The National Institute of Environmental Health Sciences (T32 ES007060‐31A1) provided software used for figure construction.

### Author Declarations

This research was approved as exempt by the Institutional Review Boards of the University of New England and Geisinger.

